# Enhanced Recovery after Intensive Care (ERIC): study protocol for a stepped-wedge cluster randomized controlled trial to evaluate the effectiveness of a critical care telehealth program on process quality and functional outcomes

**DOI:** 10.1101/2020.01.25.19013565

**Authors:** Christine Adrion, Björn Weiss, Nicolas Paul, Elke Berger, Reinhard Busse, Ursula Marschall, Jörg Caumanns, Simone Rosseau, Ulrich Mansmann, Claudia D. Spies, on behalf of the *ERIC* study group

## Abstract

**Introduction:** Survival after critical illness has noticeably improved over the last decades due to advances in critical care medicine. Besides, there are an increasing number of elderly patients with chronic diseases being treated in the intensive care unit (ICU). More than half of the survivors of critical illness suffer from medium- or long-term cognitive, psychological and/or physical impairments after ICU discharge, which is recognized as post intensive care syndrome (PICS). There are evidence- and consensus-based quality indicators (QIs) in intensive care medicine, which have a positive influence on patients’ long-term outcomes if adhered to.

**Methods and analysis:** The protocol of a multicentre, pragmatic, stepped wedge cluster-randomized controlled, quality improvement trial is presented. During three predefined steps, 12 academic hospitals in Berlin and Brandenburg, Germany, are randomly selected to move in a 1-way crossover from the control to the intervention condition. After a multifactorial training programme on QIs and clinical outcomes for site personnel ICUs will receive an adapted, interprofessional protocol for a complex telehealth intervention comprising of daily telemedical rounds at ICU. The targeted sample size is 1431 patients. The primary objective of this trial is to evaluate the effectiveness of the intervention on the adherence to 8 QIs daily measured during the patient’s ICU stay, compared to standard of care. Furthermore, the impact on long-term recovery such as PICS-related patient-centred outcomes including health-related quality-of-life, mental health, clinical assessments of cognition and physical function, all-cause mortality, and cost-effectiveness 3 and 6 months after ICU discharge will be evaluated.

**Ethics and dissemination:** This protocol was approved by the ethics committee of the Charité Universitätsmedizin, Berlin, Germany (EA1/006/18). The results will be published in a peer-reviewed journal and presented at conferences. Study findings will also be disseminated via the website (https://www.eric-projekt.de).

**Trial registration number:** ClinicalTrials.gov NCT03671447 (https://clinicaltrials.gov/ct2/show/NCT03671447, 22 August 2018)

**ARTICLE SUMMARY:** Strengths and limitations of this study

▪ Telemedicine-based care potentially improves the adherence to quality indicators (QIs) in intensive care medicine, which accelerate patient recovery and improve long-term outcomes after critical illness.
▪ ERIC is the first large-scale cluster-randomized controlled trial to be carried out in ICUs in Berlin and Brandenburg, Germany, comparing the clinical and cost effectiveness of a telehealth-based quality improvement intervention to standard of care.
▪ By employing a stepped-wedge design, this quality improvement study will allow each cluster to act as its own control and preserve the internal validity of the study, with a potential for confounding by secular trends.
▪ The nature of the intervention does not allow blinding of study personnel and eligible patients at ICUs and might be confronted with cross-contamination and staff turnover.
▪ ERIC allows getting a comprehensive evaluation from the patient’s perspective, healthcare staff and health economics and assessing its suitability to become standard of care.

## INTRODUCTION

There is substantial heterogeneity in the process of critical care worldwide.[1] With more than 2.1 million critical care cases (25/1000) per year, Germany has one of the most dense critical care environments in developed countries.[2] Over the last two decades, increased life-expectancy, demographic changes and progress in treatment have resulted in increased survival, thus, resulting in trajectories typical for this new cohort of ICU survivors.[3]

These trajectories are characterized by impairments including mental illness (i.e. anxiety, post-traumatic stress disorder (PTSD), depressions), neurocognitive degeneration and neuromuscular end-organ failure resulting in conditions such as long-term ventilation. These long-term consequences are summarized as Post-Intensive Care Syndrome (PICS) [4, 5] and affect a considerable number of ICU patients. For example, 40 % of mechanically ventilated ICU survivors have measurable cognitive impairments three months post discharge. More than half of these impairments are comparable to a mild and moderate Alzheimer’s dementia. The majority of these cognitive impairments persist after one year.[6] Anxiety affects half of the patients after ICU with 7% to 17% suffering from symptoms of post-traumatic stress.[7-9] The neuromuscular system is affected in 48% to 96% of ICU survivors, which reduces the walking-distance significantly even 5 years after critical illness.[10] Aside from PICS, there maintains a sub-cohort of ICU patients showing ‘chronic critical illness’ (CCI), characterized as a state of chronic dependence on organ support. Nowadays, these patients receive care in very heterogeneous settings, from rehabilitation centers to long-term acute care facilities or nursing homes.

There are evidence-based strategies to reduce the PICS/CCI burden. These include, for example, the prevention of delirium, the preference of no or light sedation over heavy sedation, the conduction of spontaneous breathing trials for timely liberation from the ventilator, and the use of quality indicators. The German Interdisciplinary Association of Intensive Care and Emergency Medicine (DIVI) has summarized quality indicators for intensive care treatment as the ‘German Quality Indicators of Intensive Care’ (QIs), with the first version established in 2010.[11] QIs give a framework for procedural as well as structural measures to improve patient outcomes and align healthcare resources, enabling their operationalization and measurement in the health system and beyond intersectoral barriers. Yet, the adherence to QIs is comparatively low, and comprehensive implementation strategies are often lacking.[12, 13]

Telemedicine in critical care can be used as a vehicle to transport content and quality to medical settings and has already become a cornerstone in care settings with reduced access to intensive care specialists.[14, 15] It typically consists of a telemedical cockpit and a remote site, which share an audio-visual connection and health data to various degrees (depending on the applied care model).[16] Several studies and meta-analyses revealed a significant reduction in ICU mortality and lengths of stay at ICU by telemedicine applications.[17-19] Nevertheless, data on alternative aspects of effectiveness of ICU telemedicine programs are limited and conflicting.[20, 21] Building on this evidence, there exists to our best knowledge no randomized controlled trial investigating whether a virtual care-network is capable of increasing quality of care and decreasing functional impairment of ICU survivors.

To fill this evidence gap, the quality improvement trial *ERIC* (Enhanced Recovery after Intensive Care) was initiated. This project is funded by the German Innovation Fund (‘New Forms of Care’) coordinated by the Innovation Committee of the Federal Joint Committee (grant number 01NVF16011; https://www.g-ba.de/english/).[22]

## METHODS AND ANALYSIS

This trial protocol is presented in accordance with the SPIRIT Statement [23, 24] and also considers the CONSORT reporting guideline for stepped wedge cluster randomised trials (SW-CRT) [25, 26], the Standards for Quality Improvement Reporting Excellence (SQUIRE) reporting guideline [27] and the TIDieR checklist [28]. The project’s webpage (in German) provides an overview for clinicians, patients and their relatives and can be accessed at https://www.eric-projekt.de.[29]

### Aim and objectives

This trial aims to demonstrate the clinical effectiveness of a targeted multifaceted quality improvement intervention mediated by a critical care telemedicine service at ICU. The project with a target recruitment goal of 1431 patients in total has the following primary objective:

▪ to evaluate the benefit of a complex behavioural and telemedicine based intervention on the adherence to evidence-based, national QIs daily assessed during the patient’s ICU stay (intra-sectoral modification).[30]

Secondary objectives are:

▪ to evaluate whether the intervention improves long-term core outcomes including overall survival, health-related quality-of-life (HRQoL), and other (PICS-related) patient-centred outcomes concerning mental health, cognition and physical function of ICU survivors 3 and 6 months post ICU discharge when compared to standard care;
▪ to estimate, in a health-economic analysis alongside the main trial, the cost-effectiveness during the 6 month post-ICU follow-up for patients exposed to the intervention versus to the control condition at ICU. We aim to assess whether the intervention imposes lower costs and care needs than with routine practice, e.g. by reducing the proportion of patients discharged ventilated from ICU.

### Study design and setting

ERIC is a national, large-scale, multicentre, pragmatic, cluster-randomized controlled trial with an open cohort stepped-wedge design with continuous recruitment.[31] The study will be conducted in adult critical care units of hospitals located in the metropolitan area of Berlin and the rural area of the surrounding federal state Brandenburg with a population of about six million inhabitants in total. The study area has approximately 150,000 ICU admissions per year (GBE-Bund health data [32]). During three predefined steps over the study period of 25 months (first-patient-in to last-patient-out) which includes a six month post-ICU follow-up at the patient level, participating hospital facilities are randomly selected to move in a 1-way crossover from the control to the multicomponent intervention condition.

### Site selection

Hospitals defined as study sites are eligible to participate if they are able to commit to the following criteria at the institutional level:

▪ Providing adult critical care units;
▪ Located in the Berlin/Brandenburg metropolitan region;
▪ Adherence to general legal obligations to participate in the study funded by the German Innovation Fund and participate in the respective contracts (which includes a cooperation agreement with the Charité – Universitätsmedizin Berlin);
▪ Adherence to cluster-randomization.

Trial sites (comprising of one to three ICUs) were recruited from the set of facilities according to their letter-of-intent written during the project application ensuring that recruitment, data collection and delivery of the telemedicine-based intervention are feasible.

### Patient population and eligibility criteria

Patients admitted to the ICU at the participating site will be routinely screened against the following eligibility criteria:

#### Inclusion criteria at the participant level

▪ Age 18 years or greater;
▪ Expected to receive treatment in a mixed, medical or surgical ICU connected to the project for more than 24 hours;
▪ Coverage by a German statutory health insurance company;
▪ Written informed consent of patient or legal representative.

#### Exclusion criteria at the participant level

▪ Age less than 18 years;

### Patient recruitment, and informed consent model

Prior to trial commencement, participating sites have to provide informed consent on an institutional level. Due to the open cohort design patients are recruited in continuous time as they become eligible, i.e. usually at ICU admission, and most of them are exposed for a short time.[33] Patients will be identified and screened for eligibility by the local team in the ICU which has been trained by the central study coordination team of the Charité (consortium leader; for details concerning the structure of the ERIC consortium see supplementary file 1). Written informed consent is obtained by the patient or the legal representative if the patient is unable to consent.

### Randomization, allocation concealment and masking

Due to the stepped wedge design, hospitals defined as clusters are randomized to receive the experimental intervention at different pre-planned crossover dates (‘steps’), and all clusters receive it (Figure 1). Prior to trial commencement 12 sites that have provided a letter-of-intent to be involved in the study will be randomized. The independent trial statistician contemporaneously randomized the sites to the three sequence groups using a computer-generated algorithm (nQuery Advisor V.7). Concealment of the crossover date assignment from sites and the research team is not possible due to the inevitable planning of the preceding training period for ICU personnel. Patients will generally be aware of the condition they are exposed to. However, patients (and their proxies) will be unaware of the allocation sequence, i.e. those not yet receiving the intervention will not be aware of the time at which the intervention is implemented at the treating ICU. By nature of the trial design and the intervention, it is not possible to blind the study personnel at ICUs. After ICU discharge, healthcare providers, interviewer staff and general practitioners conducting follow-up assessments have no access to telemedical data and therefore will be kept blinded.

**Figure 1.**
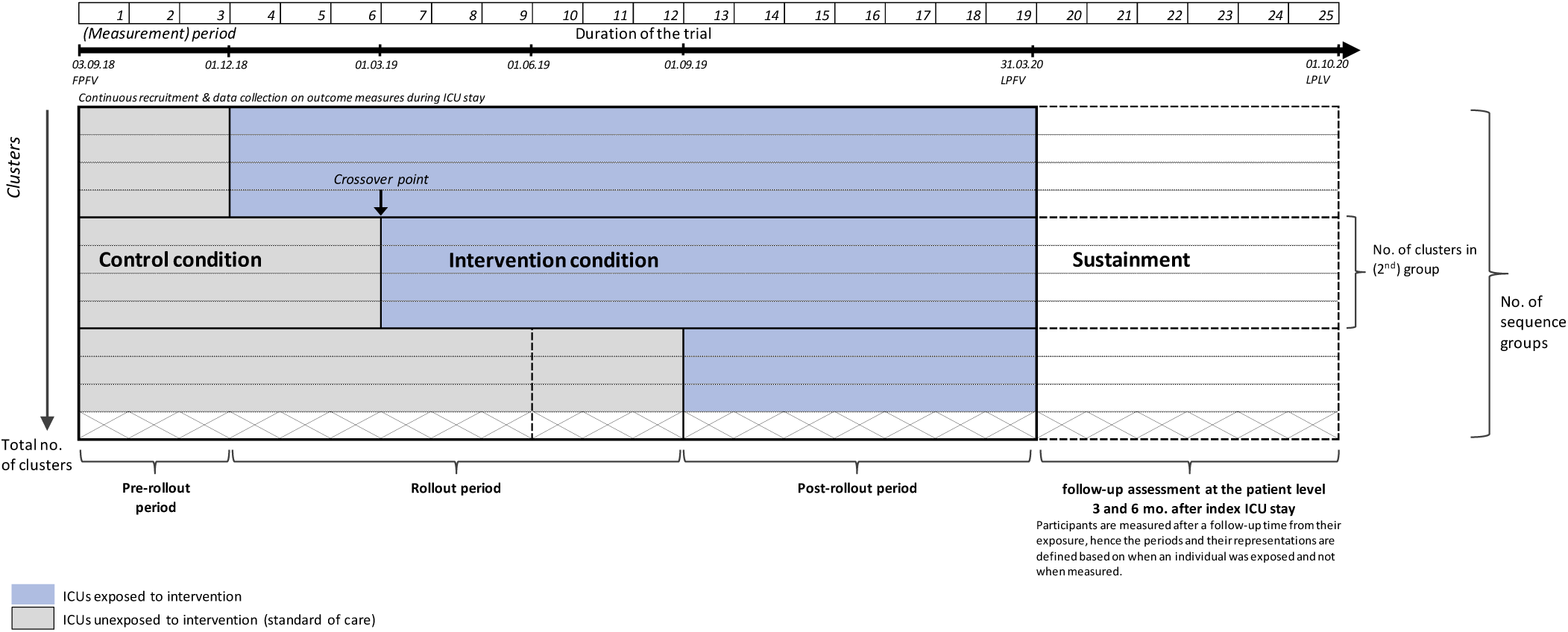
Schematic illustration of the rollout process, timeline, and study design applied in the ERIC project (after the decision to extend recruitment, including postponement of 3rd crossover date (indicated as vertical dashed line)). Blue areas indicate intervention stage (telemedicine), grey areas control stage, and the unshaded area indicates the post-ICU phase (displayed after LPFV). In each sequence group, a 3 month transition period at the end of the control condition is defined. Follow-up assessments (secondary outcomes) on the patient-level 3 and 6 months after index ICU stay. Randomization of 12 units occurred at a single time point prior to patient enrolment; one cluster of sequence group 3 withdrew IC prior to start of recruitment. (Figure adapted according to [33].)

Although several outcome assessors including data analysts will not be blinded, we do not expect a high risk of ascertainment bias to influence the treatment effect for objective outcomes.

### Treatment conditions and implementation of the intervention

Figure 2 represents a schematic representation of the pillars of the ERIC intervention.

**Figure 2.**
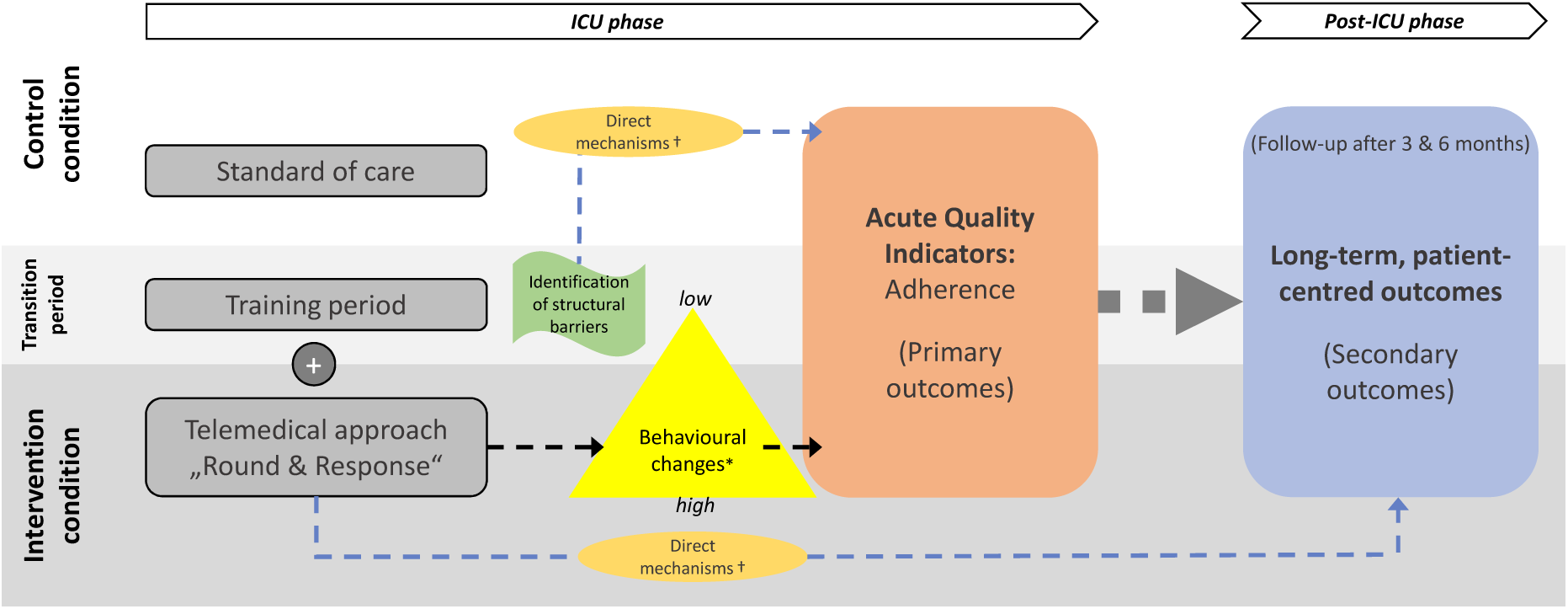
The pillars of the *ERIC* intervention: An integrated approach to critical care and causal pathways. * Behavioural changes include process-related factors: planning and coordination of measures, risk-benefit evaluation, responsibilities, and roles. Cultural barriers: lack of mobility culture, staff knowledge and clinical expertise, prioritisation. † Direct mechanisms are mechanism that might influence the QI adherence; the structural barriers might have an influence on the QI adherence. The training period will be considered within the statistical analyses.

#### Intervention condition: telemedicine

A complex health-related behavioural, quality improvement intervention comprising of the following two core components will be implemented on an institutional level:

1. Structured daily, *tele-medical cart-based* ward rounds will be conducted guided by QIs in intensive care medicine (Version 2017 published by the DIVI, definition according to [30]) in order to assess patient-individual QI performance measures. These *QI-visits* will be led by a specialised ICU-consultant and a critical care-trained nurse working in the telemedical cockpit *(tele-ICU*, located at the cluster Charité at the site of the consortium leader) and attended by the concomitant treating physician and the bedside nurse at the *remote-ICU*. This telehealth approach involves an interactive and secure two-way audio-visual communication at the bedside of the patients and face-to-face dialog during delivery of healthcare between remote and local care provider (remote video visualization of patients and their monitoring devices by means of the tele-medical cart serving as access device).
2. A *24/7 on-call service* staffed with a board-certified critical care specialist will be provided by the telemedical cockpit to ensure the coverage of acute medical issues on demand and easy access to high-quality ‘virtual care’ in the local treating ICU.

The main features of the tele-ICU and its equipment together with a photograph of the telemedical cart (manufacturer: InTouch Technologies®, Inc., USA) utilized in the local patient rooms are provided in the online supplementary file 2 following the TIDieR checklist.[28]

#### Control condition: usual ICU care

While delivering the control condition ICUs are provided with no special instruction in the care of their patients and treatment is considered to be ‘usual care’ according to local standards, i.e. the status quo provided by the cluster’s ICU before the randomized start of the intervention, with treatment at the discretion of the treating clinician. In particular, no telemedicine-based support will be provided during the daily bedside ward rounds (QI-visits) conducted to assess the QI-related parameters.

#### Preceding training activities and implementation of the intervention

A structured multi-component training program will be provided to all hospitals within a sequence group taking place during the last 3 months before the crossover date representing a transition phase. The training is delivered as a blended-learning concept with 1.) an e-learning course for each QI (ERIC e-learning platform accessible at https://best-edx.charite.de), 2.) a simulation-based training, and 3.) an on-the-job training to make sure the local ICU staff is trained in the use of the telemedical cart. Not the whole ICU team at the participating site will be trained, but local experts (physicians and nurses) can operate as multiplicators within the team.

Prior to the training period, ICUs are highly encouraged to consider a *medical peer review* as a standardized tool for continuous quality improvement in intensive care medicine which was developed by the German Medical Association (based on the conceptual framework of the Plan-Do-Study-Act (PDSA) cycle) [34]). These peer reviews conducted during mutual visits by colleagues promote the exchange of experience between professions and disciplines at ICU and focus on the systematic evaluation of the quality of an ICU’s structure, its processes and outcome to secure the sustainability of the planned change processes in patient care.[35-37]

#### Adherence to the intervention

After the crossover date, the adherence to the intervention condition delivered by the ICU is monitored by the tele-ICU team. The login-times to the audio-visual communication are monitored by a distinct fleet management system resulting in an anonymous monthly performance report and show the connection times that enable to draw conclusions about the compliance of the caregivers both at the remote as well as at the tele-ICU. Unusual durations or timings of ward rounds will be reported to the clinical lead of the tele-ICU.

### Study outcomes and data collection schedule

#### Primary Outcomes

To evaluate whether the intervention has a beneficial effect compared to the control condition in at least one of the 8 patient-level QIs in intensive care medicine (definition according to Kumpf *and colleagues* [30]), 8 co-primary binary efficacy outcome measures derived from several QI-related performance parameters are pre-specified (Table 1), with each one of these defined as follows:

**Table 1.** Intra-sectoral Quality Indicators Intensive Care for Germany (third edition 2017, see [30]) applied for the definition of the binary primary outcomes.

| Domain No. | Quality Indicators Intensive Care |
| --- | --- |
| **QI I** | Daily multiprofessional and interdisciplinary clinical visits with documentation of daily goals |
| **QI II** | Management of sedation, analgesia, and delirium |
| **QI III** | Patient-adapted ventilation |
| **QI IV** | Early weaning from invasive ventilation |
| **QI V \*** | *Monitoring the measures for the prevention of infection* |
| **QI VI** | Measures for infection management |
| **QI VII** | Early enteral nutrition |
| **QI VIII** | Documentation of structured patient and family communications |
| **QI IX** | Early mobilization |
| **QI X \*** | *Direction of the intensive care unit* |

\* QI V and QI X are not specified as primary efficacy outcomes since they are not assessed on a patient-level during daily QI visits, and do not enable estimation of an interpretable effect of the intervention.

▪ Adherence [fulfilled yes/no] to a single intra-hospital QI being daily assessed on a patient-level starting from date of enrolment (after ICU admission) until ICU discharge, within a 24 hour time window.

Whether a QI for patient *i* on day *t* is fulfilled or not is subsequently assessed by an intensive care specialist at the tele-ICU cockpit rating the underlying electronic QI documentation recorded by the remote-site physician. This central endpoint adjudication process is utilized irrespective of whether the ICU delivers care on control or intervention condition.

#### Secondary Outcomes

Secondary outcomes will be assessed during the sustainment phase with time points scheduled after a 3 and 6 month follow-up time from the patient’s exposure during index ICU stay. In particular, several key secondary outcomes are defined following the conceptual framework of core outcome sets with respect to PICS-related domains.[5, 38] Besides, some measures related to healthcare utilization and socioeconomic status will be assessed.

▪ All-cause mortality up to 6 months following the first study-related ICU admission: The number of deaths from any cause including in-hospital mortality will be recorded (using hospital administrative records, electronic medical records, municipal personal records database and the 3- and 6-month follow-up with surrogates);
▪ Mental health condition (domains: anxiety and depression screen) at month 3 and 6: The patient-reported symptom burden on anxiety and depression will be assessed by the paper-based Patient-Health-Questionnaire PHQ-4. Higher total scores indicate higher impairment;[39]
▪ Mental health condition (domain: PTSD) at month 6: The patient-reported symptom burden on post-traumatic stress evaluated using the paper-based questionnaire Impact of Event Scale Revised (IES-R). Higher total scores indicate greater distress;[40]
▪ Cognition (domains: memory, visuospatial and visuoconstructional skills) at month 3 and 6: Functional outcome as assessed by the MiniCog test (2 tests: three-item recall task; clock-drawing task). Higher scores indicate better cognitive functioning;[41, 42]
▪ Cognition (domain: verbal fluency) at month 3 and 6: Functional outcome as assessed by the Animal Naming Test. Higher scores indicate better cognitive functioning;[43]
▪ Physical Function at month 3 and 6: The patient’s physical function, walking ability and risk of fall as assessed by the Timed Up & Go (TUG) test. Higher scores indicate a higher level of impairment;[44]
▪ Physical Function at month 3 and 6: The patient’s muscle and nerve function assessed by the Hand-Grip-Strength (HGS) test is measured with a dynamometer (average strength [in kg] of three trials for the dominant hand);[45]
▪ HRQoL at month 3 and 6: Patient’s self-reported HRQoL as measured by the EuroQol - 5 Dimensions - 5 Level (EQ-5D-5L) descriptive system and the Visual Analogue Scale (VAS). The EQ-VAS is a thermometer-like rating scale ranging from 0 (worst imaginable health state) to 100 (best imaginable health state);[46, 47]
▪ Organ dysfunction at month 3 and 6: number of patients with organ dysfunction as assessed by GP or study personnel/ investigator;
▪ Pulmonary Function and symptoms – Dyspnea at month 6: self-perceived breathlessness during daily activities graded by the Modified British Medical Research Council (mMRC) Dyspnea Scale [range 0 to 4], with a higher score indicating higher impairment;[48, 49]
▪ Outpatient ventilation: total duration [in days] of mechanical ventilation up to 6 months after index ICU discharge;
▪ Functioning and Disability at month 6: patient-reported general disability score, as measured by the WHO Disability Assessment Schedule (WHODAS 2.0) for activity limitation and participation restriction, 12-item short version, self-administered questionnaire. The raw score is calculated by summing the values for each item. Higher scores indicate greater disability;[50, 51]
▪ Length of stay (LOS) at ICU, and at hospital: Total number of days spent in ICU / in a hospital, as assessed at month 6;
▪ Employment status (including e.g., return to work or change in employment status) at 3 and 6 months;

#### Data collection and trial procedures during ICU stay and follow-up

Table 2 shows the patient data collection.

**Table 2.**
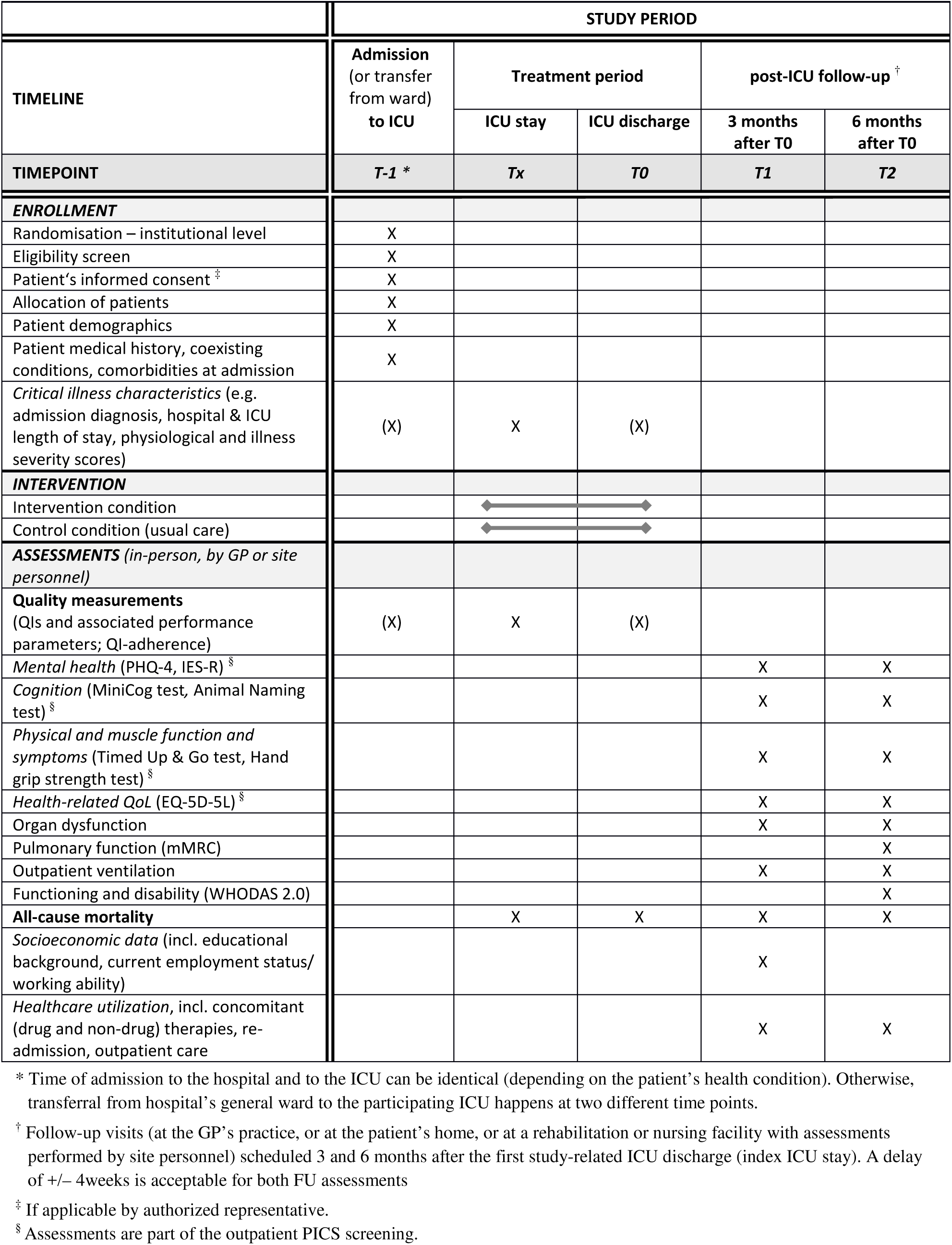
Schedule of enrolment, interventions and assessments in the ERIC trial from the patient’s perspective.

|  | STUDY PERIOD |  |  |  |  |
| --- | --- | --- | --- | --- | --- |
| TIMELINE | Admission<br>(or transfer<br>from ward)<br>to ICU | Treatment period |  | post-ICU follow-up<sup>†</sup> |  |
|  |  | ICU stay | ICU discharge | 3 months<br>after T0 | 6 months<br>after T0 |
| TIMEPOINT | T-1 * | Tx | T0 | T1 | T2 |
| **ENROLLMENT** |  |  |  |  |  |
| Randomisation – institutional level | X |  |  |  |  |
| Eligibility screen | X |  |  |  |  |
| Patient's informed consent<sup>‡</sup> | X |  |  |  |  |
| Allocation of patients | X |  |  |  |  |
| Patient demographics | X |  |  |  |  |
| Patient medical history, coexisting conditions, comorbidities at admission | X |  |  |  |  |
| Critical illness characteristics (e.g. admission diagnosis, hospital & ICU length of stay, physiological and illness severity scores) | (X) | X | (X) |  |  |
| **INTERVENTION** |  |  |  |  |  |
| Intervention condition |  |  |  |  |  |
| Control condition (usual care) |  |  |  |  |  |
| **ASSESSMENTS** (in-person, by GP or site personnel) |  |  |  |  |  |
| Quality measurements (QIs and associated performance parameters; QI-adherence) | (X) | X | (X) |  |  |
| Mental health (PHQ-4, IES-R)<sup>§</sup> |  |  |  | X | X |
| Cognition (MiniCog test, Animal Naming test)<sup>§</sup> |  |  |  | X | X |
| Physical and muscle function and symptoms (Timed Up & Go test, Hand grip strength test)<sup>§</sup> |  |  |  | X | X |
| Health-related QoL (EQ-5D-5L)<sup>§</sup> |  |  |  | X | X |
| Organ dysfunction |  |  |  | X | X |
| Pulmonary function (mMRC) |  |  |  |  | X |
| Outpatient ventilation |  |  |  | X | X |
| Functioning and disability (WHODAS 2.0) |  |  |  |  | X |
| All-cause mortality |  | X | X | X | X |
| Socioeconomic data (incl. educational background, current employment status/working ability) |  |  |  | X |  |
| Healthcare utilization, incl. concomitant (drug and non-drug) therapies, re-admission, outpatient care |  |  |  | X | X |

\* Time of admission to the hospital and to the ICU can be identical (depending on the patient's health condition). Otherwise, transferral from hospital's general ward to the participating ICU happens at two different time points.
<sup>†</sup> Follow-up visits (at the GP's practice, or at the patient's home, or at a rehabilitation or nursing facility with assessments performed by site personnel) scheduled 3 and 6 months after the first study-related ICU discharge (index ICU stay). A delay of +/- 4 weeks is acceptable for both FU assessments
<sup>‡</sup> If applicable by authorized representative.
<sup>§</sup> Assessments are part of the outpatient PICS screening.

#### ICU stay

Data documented by local research teams for all patients while at ICU includes:

▪ patient details (distinct identifiers, socio-demographics, statutory health insurance status)
▪ baseline data and critical illness characteristics (e.g. date and mode of hospital or ICU admission or transfer from general ward care, eligibility criteria, vital signs, laboratory findings, documented pre-existing conditions/medical history, illness severity scores)
▪ QI-related performance parameters assessed during daily QI-visits
▪ Hospital discharge data (discharge status (intermediate care/normal care/death), date of discharge/death)

#### Follow-up procedures and post-acute care

In providing written informed consent, all patients are agreeing for the trial team to have access to their medical records for data collection and to being contacted in order to arrange two follow-up assessments. These are scheduled 3 and 6 months after index ICU discharge including an enhanced patient monitoring with the opportunity for on-demand counselling sessions between the treating GP or the rehabilitation facility and investigators of the Charité experienced in ICU aftercare including PICS. Locating of patients will contemporaneously be supported by contacting the patient’s GP starting after one month, providing information about the ERIC project and relevant follow-up procedures. If no GP is available or if the patient’s GP does not support the project, site personnel affiliated to the Charité (consortium leader) or to a consortium partner (hospital EvB Bad Belzig) will conduct the follow-up assessments. The latter one is specialized for out-of-hospital mechanical ventilation and prolonged weaning in the case the patient stays at a rehabilitation facility, weaning unit or a hospital at the time of the follow-up visits. Depending on the patient’s health status, follow-up visits may be performed as home visits by clinicians of the study team.

Post-ICU follow-up data include health service and resource use assessed by means of medical records and will be documented by paper-based CRFs delivered to the patient’s GP or filled by the study team. Additionally, validated tests and patient questionnaires with a focus to screen for PICS-related symptoms will be used (see Table 2 for measurement instruments). Survival status at 3 and 6 months will be ascertained through the patient’s surrogates, the GP or caregivers, or municipal personal records database.

### Data management, data security and quality control

Patient-level study data documented during the ICU stay will be collected and managed using REDCap® (Research Electronic Data Capture) hosted at Charité at the site of the coordinating investigator. REDCap is a secure, web-based research data management platform providing 1) an intuitive interface for validated data entry; 2) audit trails for tracking data manipulation and export procedures; 3) automated export procedures for seamless data downloads to common statistical packages; and 4) procedures for importing data from external sources.[52] Follow-up data at month 3 and 6 are initially collected via paper-based CRFs and questionnaires before being entered into a separate REDCap CRF. All installations have been made in compliance with EU GDPR and are continuously monitored by the data protection officers.

Data quality control will be assured by automated data entry plausibility checks, on-site monitoring to ensure accuracy and enquire implausible or missing data on a regular basis.

### Patient and public involvement

No patients were involved in setting the research question or the outcome measures, nor were they involved in developing plans for participant recruitment, or the design and implementation of the study. A patient representative is a member of the trial oversight committee (independent advisory board). Public involvement is achieved through the active role of the BARMER statutory health insurance company that represents the interests of its members. The results of the study will be disseminated to the patients through public information such as the BARMER customer magazine and by the dedicated project’s website which has a section for patients and relatives. Furthermore, our aim is to include patients in the interpretation of the study results if possible.

### Clinical evaluation: Statistical methodology and planned analyses

#### Power considerations and pre-trial sample size calculation

Following the cluster-randomized design, a pragmatic power calculation was performed on a per hospital facility basis assuming equal cluster sizes and considering a limited number of potential sites and resources. To allow for eight binary co-primary outcomes with all of them having equal importance from a clinical perspective, a Bonferroni correction for multiple testing was applied for sample size calculation based on an overall one-sided type I error rate of alpha = 5% (resulting in an alpha/8 = 0.625% significance level for confirmatory testing for a single QI). A minimum clinically relevant difference of 10% in QI adherence was specified; this target difference between both treatment conditions was guided by values in the literature.[12, 53]

A two group χ^2^ test with a 0.625% one-sided significance level will have 82% power to detect the difference between a Group 1 proportion (control condition), *p*_1_, of 60% and a Group 2 proportion (intervention condition), *p*_2_, of 70% (odds ratio of 1.556) when the sample size on each treatment condition is 530 patients in the case of independent observations (nQuery Advisor V7.0). To deal with the correlation between individuals from the same cluster, a design effect (variance inflation factor) of 1.35 was estimated, together with an intracluster correlation coefficient of 0.117 (derived from unpublished data available for site Charité only) which measures the correlation between observations within the same cluster.[54, 55] The total study sample size required for the CRT design is then obtained as 1431 patients. Considering practical aspects of admission and capacities results in a total setting of 12 clusters with 163 cases per cluster and a total sample size of 1956 cases (12· 163). Further details are given in the online supplementary file 3.

No transition period was included in the sample size calculation. Besides, there has been no allowance for varying cluster sizes since these methodological issues still need development for studies with stepped wedge design. Altogether, we expect the underlying assumptions used for the initial sample size calculation being rather conservative to detect an absolute increase in adherence of 10% in at least one of eight QIs.

#### Statistical methods – clinical effectiveness

The analyses of primary outcomes will be conducted according to intention-to-treat. All patients from a randomized medical facility defined as a unit of randomization (one cluster comprising of 1 to 3 ICUs) who are recorded in the database and for whom at least one QI measurement has been obtained will be included in the analyses with respect to the treatment specified by the allocated randomization order. The eight co-primary outcomes will be compared using Bonferroni-adjusted two-sided confirmatory testing at a 0.625% significance level.

Medical facilities who initially agreed to participate but subsequently withdraw before trial start date but after randomization (without recruiting any patients) will be excluded. Several protocol deviations at the cluster level may occur: Departures from the randomization at a cluster level are defined as any cluster which does not switch to the intervention at the assigned intervention implementation time point according to the randomization schedule determined prior to trial commencement. Depending on the relevance and number of the deviations, a *per protocol* analysis, which will exclude departures from the randomisation schedule (including the affected patients) and any cluster withdrawal, will be completed for the primary efficacy outcomes only.

Most patients experience either the control or intervention condition during their index ICU stay (defined as the first study-related ICU stay at one of the participating medical facilities).’Crossover patients’, i.e. patients being exposed to both conditions, should be avoided in the case they are admitted to the ICU shortly before the allocated crossover date. If a patient will be re-admitted to the ICU at a later time – documented as a new *case* – he or she will be exposed to the condition delivered at the respective time point which might be different from the one during index ICU stay.

To avoid contamination, patients who will be enrolled before the ICU’s crossover with set-up/activation of the tele-ICU will remain being treated on control condition according to the protocol. Therefore, patients who will nevertheless be exposed to both conditions due to being enrolled immediately before the ICU’s crossover will be excluded from the analyses (i.e. from the primary comparison of control and intervention condition).[31] Likewise, patients who are admitted on control and later re-admitted on intervention condition after the site’s crossover, thus, being exposed to both conditions, will also be excluded from the principal analysis.

A generalized linear mixed effects modelling (GLMM) approach is chosen that allows to model intervention-by-time interactions as well as to consider assumptions on effects regarding the transition periods. Related sensitivity analyses will be described in the upcoming statistical analysis plan (SAP). More details on the models for primary and secondary outcomes are described in the online supplementary file 3. There, we also discuss methodological issues related to death truncation and informative missings for functional patient outcomes.

Statistical analyses will be performed using the software package R version 3.6.1 or higher.[56] A full SAP will be written ahead of the final database lock.

### Process evaluation

There will be nested qualitative studies embedded within this SW-CRT to evaluate the acceptability of e-learning courses during the training phase.

### Health economic evaluation

Alongside the main trial, a health economic evaluation will be performed to assess the economic impact of the ERIC intervention compared to standard of care.[57, 58] This evaluation consists of cost-effectiveness analyses (CEA) and a cost-utility analysis (CUA),[59] and the perspective of the health-, long-term-, and retirement insurances will be taken into account.[60] Direct medical and non-medical costs as well as indirect costs and outcomes will be assessed for the ICU and post-ICU period up to 12 months (by extrapolation).[61] Further, all costs and consequences will be discounted by 3%, as recommended by the Institute for Quality and Efficiency in Healthcare (IQWiG).[62, 63] Health-economic outcomes include mortality rate, rate of long-term mechanically ventilated patients, QoL as measured by EQ-5D-5L, and quality-adjusted life years (QALYs) gained.[61, 64]

The following data sources will be used, among others, to estimate costs and selected patient-reported outcomes: clinical data collected during ICU stay, hospital claims data, statutory health insurance expenditures data (BARMER), and data captured from the CRF used for follow-up assessments. Results will be reported as mean costs, mean outcomes, and incremental cost-effectiveness ratios (ICERs), where appropriate. Robustness will be addressed in sensitivity analyses, as suggested by IQWiG.[63] ERIC will be shown to be cost-effective if costs are lower and outcomes are the same or better, or if costs are the same and outcomes are clearly better as compared standard of care.

## DISCUSSION AND PRACTICAL IMPLICATIONS

### Study impact and importance

ERIC is a German large-scale cluster randomized trial with a stepped wedge design evaluating whether the implementation of a ‘round & response’ telehealth program is effective. In doing so, it is hypothesized that daily telemedicine-based, structured ward rounds being one of the core interventional components can be a successful performance-improvement strategy not only on the institutional level (clinician-led QIs as surrogates for quality-of-care at ICUs), but also on the level of the critically ill patient (benefit on patient-centred core outcomes, in particular with respect to PICS).

Given the significant financial and personnel resources required for the installation and upkeep of telemedicine systems at ICUs, a thorough evaluation of the impact of a tele-ICU coverage leading to improved intensivist coverage at off-site hospitals is vital. Assessing the process quality at ICU imposes the risk of an inadequate choice of QIs. The QIs chosen for this trial were established by the DIVI in 2010, and are evidence-based, clinical-practice guideline derived and operationalized ensuring that they can be measured on a daily basis. Eight (out of ten) consensus-based QIs are specified as binary primary outcomes which can be reliably assessed on the patient-level. However, this results in eight possible trial outcomes increasing the possibility of an equivocal rather than a definitive result.

PICS-related patient-centred outcomes such as quality-of-life will be assessed 3 and 6 months but not at baseline, that is, before ICU admission. However, given the large sample size, we assume that randomisation will balance the baseline levels of these secondary outcomes.

## Conclusion

ERIC is one of the first projects of the German Innovation Fund’s ‘New Forms of Care’ program. The project was assessed as evidence-based and regionally viable. The evaluation concept is robust and has been developed with clinicians, biometricians and health economists. It will allow a comprehensive assessment from the patient’s, clinical and health-economic perspective after about three years. If this trial demonstrates a beneficial impact on evidence-based QIs at ICU, alongside a favourable health-economic assessment, then there would be a strong case for incorporating this telemedicine program into clinical routine throughout Germany – leading to a system change in critical care medicine by improving patient care pathways.

## TRIAL STATUS

At the time of first manuscript submission, research ethics approval has been obtained for the trial. Data collection with enrolment of the first patient commenced on 04 September 2018. Last patient last visit (including a 6-month follow-up period) is expected in October 2020.

### Progress of the study and extension of the study duration

Before the transition of the last sequence group from control to intervention status we realized that the pre-planned target sample size could not be reached since the recruitment rate was far lower than anticipated. Additionally, barriers with respect to data protection rules were identified leading to a delayed cooperation agreement between several participating sites and the consortium leader. This in particular affected the number of patients recruited under control condition. One cluster of sequence group 3 withdrew informed consent prior to start of recruitment. In May 2019, all consortium partners agreed to extend the duration of the recruitment (first patient-in to last patient-in) from 12 to 19 months and postponed the pre-specified third crossover date (while extending the rollout period) by three months to further enhance the number of patients treated on control condition.

## ETHICS AND DISSEMINATION

### Ethical considerations

This study is being conducted in accordance with ethical principles that have their origin in the Declaration of Helsinki and are consistent with Good Clinical Practice (GCP). Enrolment of patients at the participating ICUs did not start until the written and unrestricted positive vote of the local ethics committee (EC) was obtained. The protocol is based on the underlying project application which received previous independent peer review as part of the grant funding process. Together with the patient information sheets and consent forms the protocol was first approved by the EC of the Charité − Universitätsmedizin Berlin on 26 January 2018 (approval number EA1/006/18), and the EC of the Brandenburg Medical School (MHB) Theodor Fontane joined (approval number Z-01-20180828). Amendments to the protocol will be submitted to the EC for review. Individual written informed consent including consent to data collection will be obtained from all eligible patients in the trial. Consent forms for the trial include consent for publication of results in peer-reviewed journals. Relevant data protection rules for all analysed data will be enforced.

By implementing a quality improvement intervention based on evidence-based QIs, no additional risks to patients are expected relative to standard of care. Outcome data are routinely collected health data together with post-ICU data. Therefore, adverse events will not be monitored or reported. An independent Advisory Board has been appointed to ensure that ethical, legal and social aspects and responsibilities are carried out according to Good Clinical Practice (GCP).

### Dissemination plan

The success of the trial will be dependent entirely on the collaboration of clinicians in the participating ICUs and those who hold key responsibilities at the study sites. The main results of the evaluation will be reported to trial collaborators and subsequently be published in peer-reviewed scientific journals and at national and international conferences. Additionally, study findings will be disseminated via a press release that will also be available on social media after publication to reach out to patients and surrogates as well as healthcare professionals. Authors and collaborators will be involved in reviewing drafts of the manuscripts, press releases and any other publication format arising from this project.

## Data Availability

Due to legal restrictions imposed by the EC and the data protection commissioner of the Charité - Universitätsmedizin Berlin, public sharing of individual deidentified participant data with other researchers or entities is not allowed. Requests may be sent to.

## Abbreviations

(S)AE(s): (serious) adverse event(s)
(e)CRF: (electronic) Case report form
CCI: Chronic critical illness
DIVI: German Interdisciplinary Society for Intensive Care Medicine
EDC: Electronic data capture
EQ-5D-5L: EuroQol 5 dimensions and 5 level version
EQ-VAS: EuroQol Visual analogue scale
EC: ethics committee
ERIC: Enhanced Recovery after Intensive Care
FPF(L)V: first patient first (last) visit
G-BA: Federal Joint Committee
GDPR: General Data Protection Regulation
GP: General Practitioner
HGS: Hand grip strength test
ICH-GCP: International conference for harmonisation (of technical requirements for pharmaceuticals for human use) − good clinical practice guideline
ICU: Intensive care unit
IES-R: Impact of Event Scale revised
ITT: Intention to treat
LPF(L)V: last patient first (last) visit
mMRC: Modified British Medical Research Council (Dyspnea Scale)
PHQ: Patient health questionnaire
PICS: Post intensive care syndrome
PP: per protocol
PTSD: Post-traumatic stress disorder
QI: Quality indicator
QALY: Quality-adjusted life year
QoL: Quality of life
REDCap: Research Electronic Data Capture
SHI: Statutory Health Insurance
SPIRIT: Standard Protocol Items: Recommendations for Interventional Trials
SW-CRT: Stepped wedge cluster randomized trial
Tele-ICU: telemedicine intensive care unit
TIDieR: Template for Intervention Description and Replication checklist
TUG: Timed Up and Go test
VAS: Visual analogue scale
WHODAS: WHO Disability Assessment Schedule

## DECLARATIONS

### Availability of data and materials

Due to legal restrictions imposed by the EC and the data protection commissioner of the Charité – Universitätsmedizin Berlin, public sharing of individual deidentified participant data with other researchers or entities is not allowed. Requests may be sent to.

### Authors contributions

CS, BW and SR did the clinical conceptual planning. UM designed the trial and has oversight for the statistical analyses; EB and RB designed the health-economic evaluation; RB has oversight for the health-economic evaluation. BW, UM and RB contributed to the preceding research proposal leading to the funding of the trial. UMar supervises the project from the perspective of the statutory health insurance company (BARMER). CS led the grant application and, as principal investigator and consortium leader, has oversight for the trial. BW is the study project manager, responsible for the study’s quality assessment and is in charge of the overall study management. CA and BW drafted the manuscript (shared first authorship). The corresponding author attests that all listed authors meet authorship criteria and that no others meeting the criteria have been omitted. All authors have critically read, contributed with inputs and revisions, and approved the final manuscript.

## Acknowledgements

We are very grateful to the clinical staff of the participating sites involved in the set-up, recruitment and data acquisition (see online supplementary file 1).

We would like to acknowledge the patients and their caregiving relatives (person of trust) or legal guardians for their support. We also thank the patient’s general practitioner’s willingness to participate in the ERIC trial during the post-ICU follow-up, and Ben Kraufmann (FOKUS, Berlin) for his ongoing advice and IT support. Further, we would like to thank the following members of the independent Advisory Board:

Christian Apfelbacher (University of Magdeburg), Martin Gersch (FU Berlin); Guenther Jonitz (Chamber of Physicians, Berlin), Cordula Mühr (patient representative of the Federal Joint Committee), Odette Wegwarth (Max Planck Institute for Human Development), Johannes Wimmer (healthcare media expert).

## Patient consent for publication

Not required.

## Ethics approval

Ethical approval for the protocol has been granted from each of the participating medical facilities involved in this trial prior to patient recruitment (leading ethics committee: ethics committee of the Charité − Universitätsmedizin Berlin; reference number EA1/006/18). This paper presents the protocol version 1.1, 27.05.2019.

## SUPPLEMENTARY DATA

**Supplementary file 1. Full list of *ERIC* study group members**

**Supplementary file 2. Detailed description of the *ERIC* telemedicine-based intervention** following the TIDieR (Template for Intervention Description and Replication) Checklist

**Supplementary file 3. Statistical Methods – further details**

**Supplementary file 4. SPIRIT 2013 Checklist**: Recommended items to address in a clinical trial protocol and related documents.

